# AKI-twinX: explainable organ structured digital twin for sepsis AKI trajectory forecasting

**DOI:** 10.64898/2026.03.13.26346736

**Authors:** Jinjin Cai, Allison E Gatz, Jiangqiong Li, Deborupa Pal, Haixu Tang, Michael T Eadon, Baijian Yang, Lingzhong Meng, Jing Su

## Abstract

Acute kidney injury in sepsis evolves over hours to days, yet most ICU models emphasize onset and provide limited insight into cardio-renal interactions. We developed AKI-twinX, an organ-structured, explainable digital twin that jointly forecasts acute kidney injury onset, acute kidney injury trajectory, and near-term mortality risk. The model learns renal and cardiovascular latent states with sparse feature gating and captures cross-organ coupling with attention. We trained AKI-twinX on MIMIC-IV sepsis using 5-fold cross-validation and evaluated it on an Indiana University Health cohort. Discrimination was consistent across systems (AUC: mortality 0.86-0.88, acute kidney injury onset 0.78-0.82, acute kidney injury trajectory 0.73-0.78). In vasopressor-treated windows, 12-hour systolic blood pressure forecasts tracked observed values (mean absolute error 8.5 mmHg). Counterfactual vasopressor withdrawal shifted predicted blood pressure downward and increased predicted risk, supporting sensitivity to clinically meaningful interventions. AKI-twinX enables trajectory-aware forecasting with bedside auditability in sepsis.

## Introduction

The concept of digital twinning, originally developed in the industrial domain, refers to the creation of a dynamic virtual replica of a physical entity or process that enables real-time interaction, simulation, and optimization.^1,2^ In medicine, digital twins are envisioned as individualized, continuously updated, and interactively linked virtual representations of patients and their therapies, forming patient–therapy “dyads” that support adaptive decision-making, risk prediction, and intervention planning,^3–6^ especially in intensive care settings.^7^ Despite this potential, most current digital twin efforts in healthcare rely on black-box models or neural policies lacking organ-specific physiological structure, making their internal reasoning difficult to interpret.^8–11^ Many are developed for oncology or chronic disease settings^9,12^ and do not target the rapidly evolving, multi-organ dynamics of intensive care, thus lacking the fine-grained temporal modeling needed to capture rapidly changing physiological states and intervention effects. Few frameworks provide transparent mechanisms for explaining how clinical features influence predictions, an essential requirement for clinical trust and adoption. Developing organ-specific, temporally resolved, and interpretable digital twins represents an important unmet need for advancing precision and real-time decision support in critical care.

A few studies have developed mechanistic, organ-specific digital twins, for example, by combining agent-based modeling, discrete event simulation, and Bayesian networks to represent multi-organ interactions in sepsis.^13^ These models offer high interpretability through expert-defined rules and causal graphs, and incorporate temporally resolved simulation of patient trajectories. However, they rely on static expert knowledge and have been tested only on small cohorts, rather than learning from large-scale real-world data, which may limit their adaptability to diverse clinical trajectories.

A key challenge in advancing physiologically grounded digital twins is ensuring transparent, clinically meaningful model interpretation. Explainable AI techniques are increasingly used in clinical modeling to enhance transparency and trustworthiness.^14^ Current approaches predominantly rely on post-hoc, model-agnostic methods (e.g., SHAP),^14,15^ ^16^ and visualization techniques such as feature or saliency maps, which are detached from model training and often provide unstable, non-physiological explanations in temporal, high-dimensional ICU data. This lack of physiologically grounded, model-integrated interpretability limits their clinical usefulness, particularly in high-stakes, real-time decision-support settings such as intensive care units. New approaches are needed to provide physiologically meaningful explanations learned jointly with model training, allowing the model itself to quantify the relative importance of different clinical features in a transparent, physiologically grounded manner. In other domains, learnable attention or gating mechanisms have been successfully applied to temporal and tabular data to identify salient features and improve interpretability.^17^

Disentangled representation learning offers a promising approach to decomposing independent generative factors, thereby improving interpretability and modularity in complex data. Yet, most existing medical applications remain abstract, lack explicit physiological structure,^18,19^ and do not disentangle organ-specific dynamics. In clinical contexts, however, structuring model representations to align with real physiological systems, such as separating cardiovascular, renal, and other organ functions, is essential for interpretability. Organ-level disentanglement provides explanations that are both physiologically meaningful and familiar to physicians, thereby enhancing clinical trust and decision support. This gap highlights the need for methods that disentangle organ-specific latent processes in a physiologically meaningful way, enabling transparent, clinically relevant predictions.

Sepsis, a major condition in intensive care, is currently defined as life-threatening organ dysfunction caused by a dysregulated host response to infection.^20^ Sepsis is responsible for about one-third of in-hospital deaths in the United States, as well as an increased risk of long-term morbidity and mortality.^21^ In septic patients, it is important to monitor vital organ function to assess disease progression and risk of death. The Sequential Organ Failure Assessment (SOFA) score comprises six systems: respiratory, coagulation, liver, cardiovascular, central nervous system, and renal. Each system is graded on a scale of 0 to 4, with 4 correlating with severe dysfunction of that system.^20^ The SOFA score provides a clinically familiar framework for assessing the vital organs and their biological functions that are crucial in sepsis patients, and its individual components predict in-hospital mortality.^22^

As such, our digital twin model borrowed components of the SOFA score to assess septic patients during their ICU stay. Among the six individual components, our model focuses on two vital organs, the heart and the kidney, while the other four are combined. These two organ systems were selected for their distinct roles and their interplay in sepsis. Cessation of cardiovascular function leads to death,^23^ and the renal system is particularly vulnerable during a septic state, with up to 70% of septic patients developing acute kidney injury (AKI).^24^ AKI can contribute to the dysfunction of other organs, and this multi-organ dysfunction can be indicative of a common underlying pathology, such as sepsis.^24^ Further, there is extensive crosstalk between the cardiovascular and renal systems. Damage to one of these systems can negatively impact the other through crosstalk, known as cardiorenal syndrome, which can occur in many conditions, including sepsis.^25,26^

The management of patients with sepsis involves empiric administration of antimicrobial agents, most commonly antibiotics, within one hour of suspected sepsis.^27^ Each hour that empiric antimicrobials are delayed, the patient’s risk of mortality increases.^27^ Providers should consider multiple variables when selecting empiric antibiotic therapy, including the most likely source of infection and the likely pathogens. Furthermore, blood cultures should be drawn before antibiotic treatment to determine the exact cause of infection.^28^ Various combinations of antibiotics may be used during the patient’s course of treatment. However, some combinations, such as vancomycin and piperacillin-tazobactam, may increase the patient’s risk of developing nephrotoxicity.^29–31^ Fluid resuscitation is also important in septic patients. If, however, patients are hemodynamically unstable despite adequate resuscitation, vasopressors are often used to maintain perfusion to vital organs. Norepinephrine is widely known as the first-line vasopressor for septic shock, with second and third-line vasopressors being vasopressin and epinephrine.^27^ Despite the conventional treatment methods used in sepsis patients, there is substantial heterogeneity.^32^ As such, our digital twin model provides patient-specific treatment methods.

To our knowledge, none of the existing models developed to predict AKI occurrence in an ICU setting has been designed to predict the progression or resolution of AKI. When AKI occurs in the ICU, renal function may recover, particularly if the KDIGO AKI stage is limited to stages 1 or 2. Thus, it is imperative that clinicians understand the trajectory of a patient’s renal function.

To address these gaps, we developed AKI-twinX, an organ-structured intensive care digital twin that integrates baseline risk information with time-varying physiologic and treatment signals to jointly forecast three clinically linked endpoints in sepsis: AKI onset, AKI trajectory, and short-horizon mortality risk. AKI-twinX represents renal and cardiovascular status as distinct yet interacting latent states, thereby enabling organ-aligned inspection alongside prediction. We trained the model on the MIMIC-IV sepsis cohort and independently evaluated it on the Indiana University Health electronic medical record cohort to assess its transportability across health systems.

### AKI-twinX model

Intensive care is the art of personalized, dynamic management of patients’ rapidly evolving clinical conditions across vital organ systems to minimize mortality. Six intensively interacting organ systems, as defined by the Sequential Organ Failure Assessment (SOFA) score, are frequently used to evaluate the performance of vital organs and estimate the death risk: the cardiovascular system (the heart), the failure of which is often the direct cause for death; the renal system, the failure of which is often the early sign of the deterioration of clinical conditions; and other organ systems including respiration, coagulation, liver, and central nervous systems.

The AKI-twinX model (Figure 1) provides three functions: monitors the health of the heart, kidney, and other vital organs; predicts the disease progress, such as the death risk and the acute kidney injury (AKI) progression trajectory; and infers the effects of candidate interventions on clinical outcomes as well as on the health of vital organs. Together, the AKI-twinX model is a real-time, personalized tool to support real-time clinical decisions in intensive care units. As illustrated in Figure 1A, for a specific patient at the current moment, the physical-to-virtual real-time twinning constructs a virtual patient according to current and historic clinical conditions, represents the status of vital organs in the latent space, and predicts the disease progression of candidate interventions, such as the death risk and the acute kidney injury (AKI) progression trajectory. The architecture of the model is shown in Figure 1 B. AKI-twinX is a generative AI for disease progression trends. The digital twin comprises a generative model and a prediction model. The generative component learns and represents the clinical trends at the moment of decision, as well as the clinical status of the heart, kidney, and other vital organs, by projecting the recent values of temporal features to a latent space with encoders and reconstructing the values of these features in the next time window with decoders. The prediction model infers the impact of candidate interventions on the risk of death and the AKI trajectory by incorporating the generative model’s latent representation of current clinical trends, baseline features, and current disease status.

**Figure 1.**
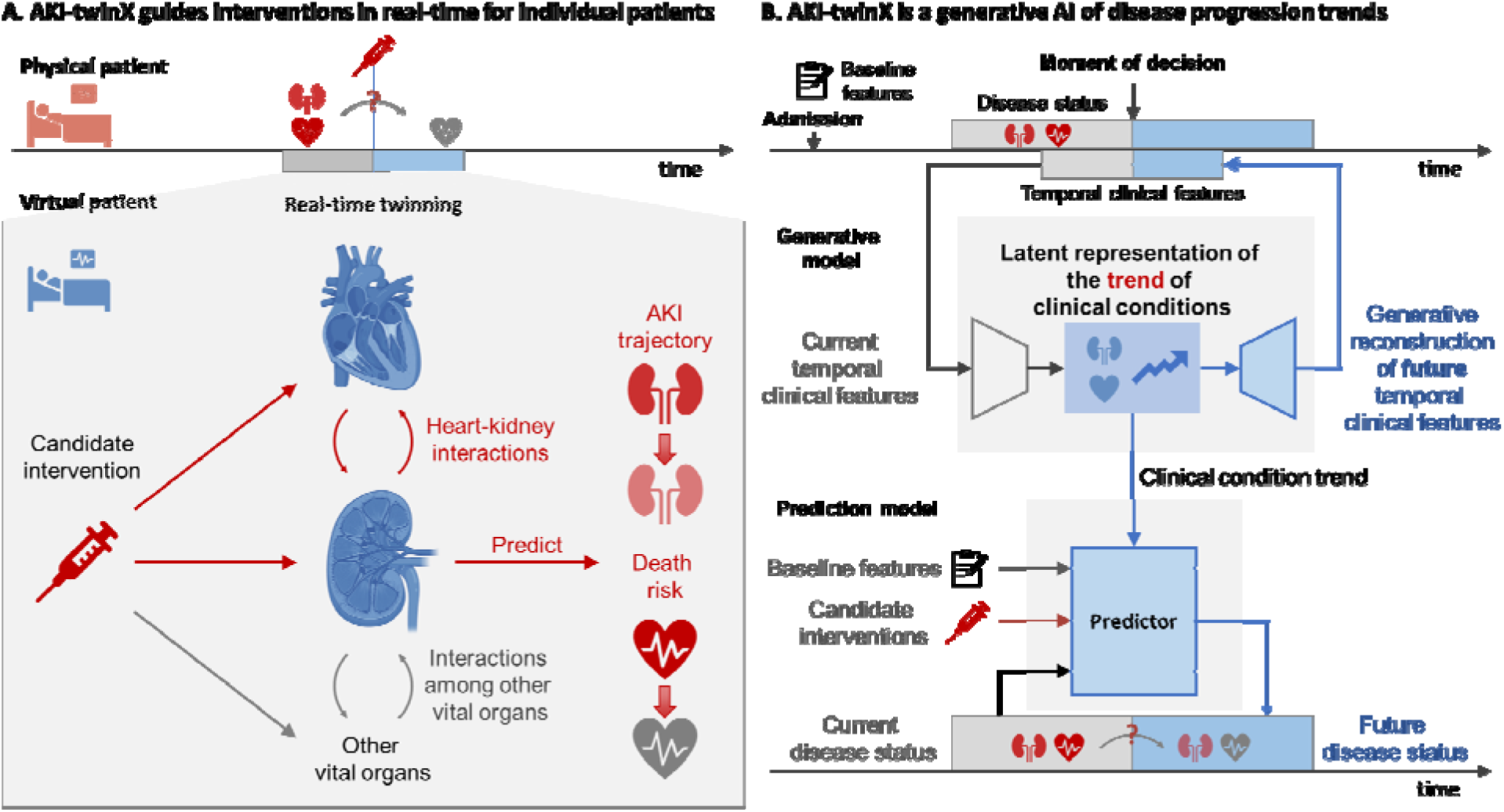
Overview of the AKI-twinX model. A. AKI-twinX guides interventions in real-time for individual patients. The digital twin model learns the latent representation of the status and clinical trends of vital organs such as the heart and kidney, infers the impact of candidate interventions on vital organs, and predicts the disease progression trends, such as the death risk and the AKI progression trajectory. B. AKI-twinX is a generative AI of disease progression trends. The digital twin is composed of a generative model and a prediction model. The generative component learns and represents the clinical trends at the moment of decision, as well as the clinical status of the heart, kidney, and other vital organs, by reconstructing the temporal clinical features in the next time window based on those in the recent time window. The prediction model infers the impact of candidate interventions on the death risk and the AKI trajectory by incorporating the latent representation of the current clinical trends from the generative model, the baseline features, and the current disease status. AKI-twinX is an explainable AI model that simultaneously learns important clinical features that

**Figure 2.**
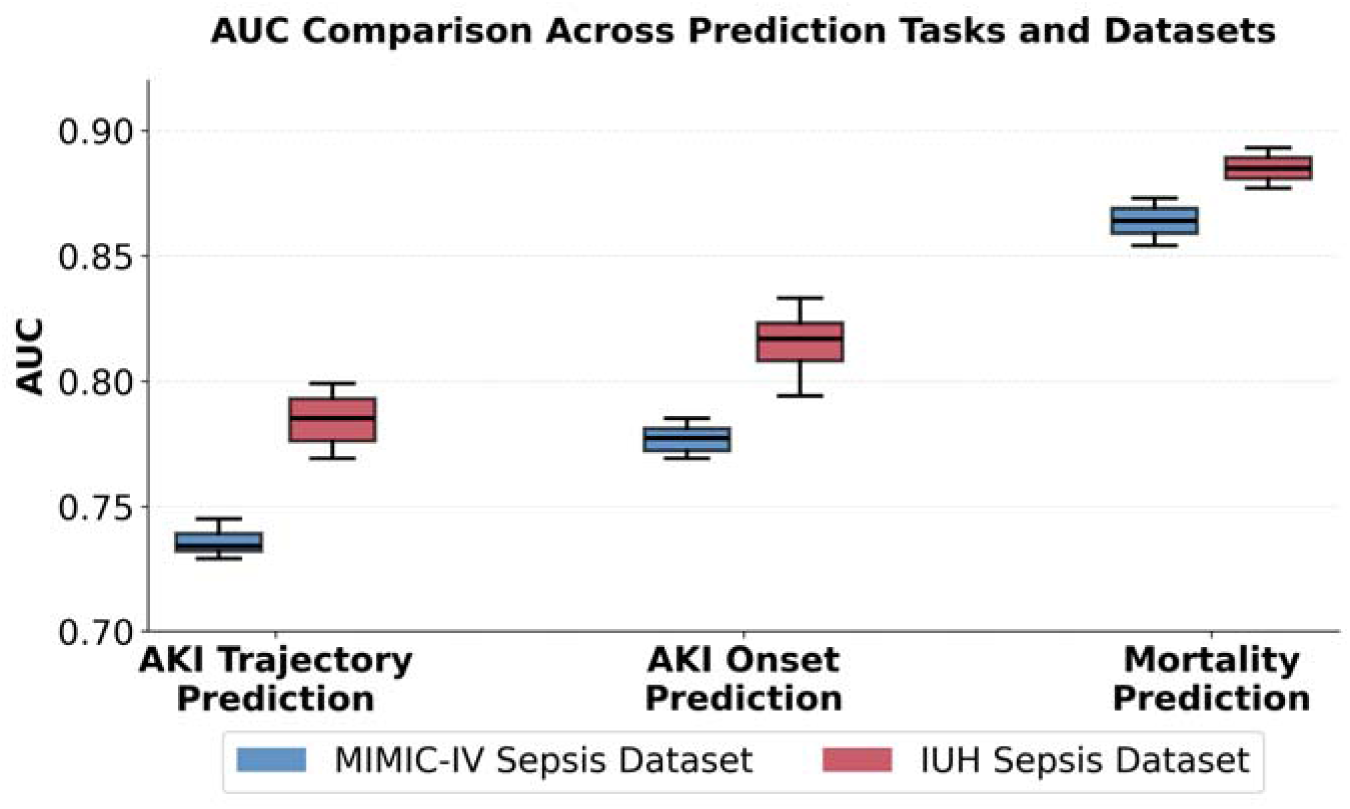
AKI-twinX performance. The model’s predictive performance is comprehensively evaluated using the predicted AKI trajectory (favorable or unfavorable), AKI onset risk, and 12-hour mortality risk. AUC: area under the receiver operating characteristic curve. AKI: acute kidney injury. IUH: Indiana University Health.

Each vital organ is distinctly represented in the latent space of the AKI-twinX model. This is achieved through organ-specific encoders and predictors, latent subspace separation between organs, organ-specific loss functions, and contrastive learning across virtual organs. For example, in the generative model, the virtual kidney is represented as a latent subspace. The virtual kidney is constructed by two kidney-specific encoders, one for temporal features and the other for baseline features. A kidney status predictor is used to predict the future renal SOFA score. An orthogonal loss is employed to ensure that information in the kidney latent subspaces is distinct from that in the rest of the latent space. Clinical information unique to the kidney status is thus extracted into the kidney latent subspace. The contribution of the virtual kidney and its interactions with other virtual organs to disease progression, such as mortality and AKI, is captured by cross-organ attention in the prediction model. Both the health of each organ and the interactions between organs can be directly visualized, supporting intuitive real-time monitoring of vital organ status and visualization of predicted changes in organ status resulting from an intervention administered or withdrawn. Specifically, the health of the heart and kidney will be presented as the predicted numerical cardiovascular and renal SOFA scores, respectively.

AKI-twinX is an explainable AI model that simultaneously learns the importance of each clinical feature for virtual organs using gating masks. This provides insights into how the digital twin predicts the effects of candidate interventions, allows clinicians to assess whether the evidence used to inform decisions is clinically meaningful, increases model transparency and trust, and generates new knowledge about important clinical features.

The selection of previous and future windows for temporal clinical features and disease progression should be determined by clinical implementation needs and data availability. We use 24-hour time windows for temporal clinical features and 48-hour time windows for death risk and AKI development, as examples.

## Results

### Model performance

The performance of AKI-twinX predictions is comprehensively evaluated by the projections of mortality risk, AKI onset risk, and AKI trajectories (favorable or unfavorable) in the next 24 hours on the MIMIC IV and the IUH sepsis cohorts. Briefly, the AKI-twinX model is trained on the MIMIC-IV sepsis cohort using a 5-fold cross-validation design. The median AUC (area under the receiver operating characteristic curve) of the prediction of 24-hour survival is 0.86 (95% confidence interval or CI: 0.85 – 0.87) on the MIMIC IV test sets and 0.88 (0.88 – 0.89) on the IU Health cohort. The AKI onset is defined as no AKI in the past 24 hours, and with AKI Stage 1 or worse in the next 24 hours. The median AUC for AKI onset prediction is 0.78 (95% CI: 0.77 – 0.97) on MIMIC-IV and 0.82 (95% CI: 0.79 – 0.84) on the IU Health cohort. The AKI trajectory is defined as the transition from the worst AKI stage in the most recent 24 hours to the next 24 hours. There are four possible AKI stages (0 – 4), yielding 16 distinct AKI stage transitions. These transitions are grouped into favorable trajectories (conditions alleviated, resolved, or stabilized, and not remaining at Stage 3) and unfavorable trajectories (conditions deteriorated or remaining at Stage 3). The median AUC of the AKI trajectory prediction is 0.73 (95% CI: 0.73 – 0.75) on MIMIC IV and 0.78 (95%CI: 0.77 – 0.80) on IU Health sepsis cohorts.

### Learned important features

The generative AI component of the AKI-twinX model learns the importance of clinical features for the heart and the kidney representations (Figure 3). Meanwhile, we performed an association analysis between clinical conditions and the cardiovascular and renal SOFA scores to facilitate interpretation of the learned feature importance.

**Figure 3.**
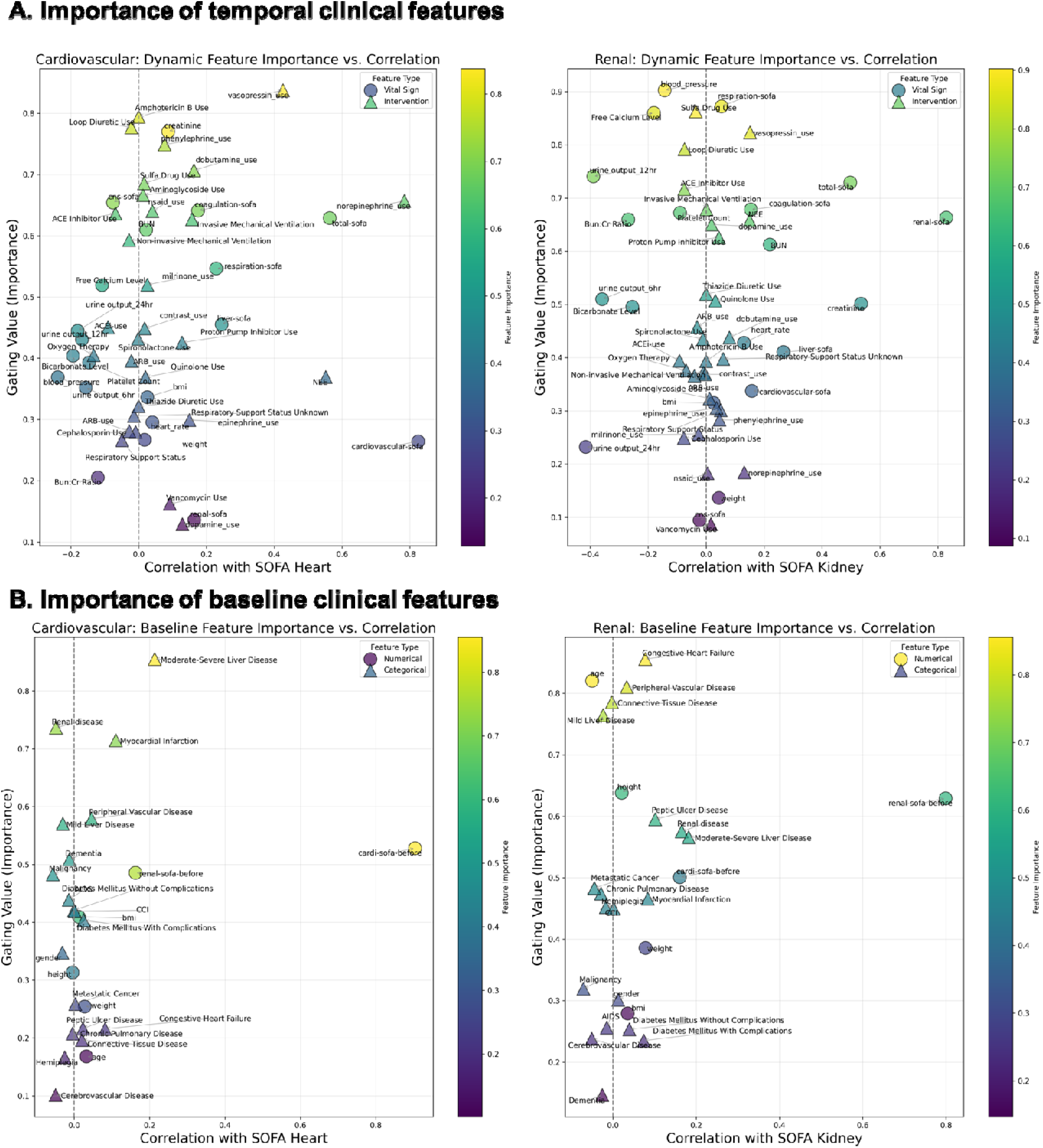
Learned feature importance for the heart and kidney status for temporal (A) and baseline (B) clinical features.

Among temporal clinical features (Figure 3A), the use of vasopressin, dobutamine, norepinephrine, and milrinone is most important, consistent with clinical knowledge. Other drugs that affect heart status include antibiotics such as amphotericin B, sulfonamides, and aminoglycosides. Other important features include mechanical ventilation, loop diuretics, non-steroidal anti-inflammatory drugs (NSAIDs), and the total central nervous system (CNS) and respiratory SOFA scores. Interestingly, kidney-related features such as serum creatinine, blood urea nitrogen (BUN), and blood free calcium level are also relevant. The most important indicator of kidney health is systolic blood pressure. Kidney status is associated with typical renal health indicators, including 12-hour and 6-hour urine output, BUN, BUN-to-creatinine ratio, serum creatinine, blood free calcium level, and the renal SOFA score. The use of vasopressin, dopamine, and the overall dose of vasopressors (summarized as the norepinephrine equivalence or NEE), loop and thiazide diuretics, sulfonamides, angiotensin-converting enzyme (ACE) inhibitors, proton pump inhibitors, and quinolone, invasive mechanical ventilation, as well as the renal, respiration, coagulation, and overall SOFA scores also significantly contribute to the inference of kidney status.

Important baseline clinical features at admission are summarized in Figure 3 B. Among them, existing liver, renal, and peripheral vascular diseases, a history of myocardial infarction, and dementia are relevant to the heart status. Age, history of congestive heart failure, preexisting peripheral vascular, liver, and renal diseases, and peptic ulcer are important for determining the kidney status.

### Importance of vasopressor use

Vasopressors such as norepinephrine, phenylephrine, vasopressin, and epinephrine are frequently used in intensive care for hemodynamic management. The use of vasopressors is crucial to maintain blood pressure in septic patients. We first demonstrate the accuracy of the predicted systolic blood pressure in patients using vasopressors (Figure 4A). The Pearson correlation coefficient is 0.76, with a mean absolute error (MAE) of 8.52 mmHg, a root mean squared error (RMSE) of 10.79 mmHg, and 95% prediction error of 21.62 mmHg. Meanwhile, the true median 12-hour systolic blood pressure is 12.0-170.5 mmHg, with the majority of patients above 90 mmHg, suggesting successful blood pressure control in most sepsis patients. Meanwhile, AKI-twinX suggests that, if the vasopressors of these patients were withdrawn, a significant drop in the systolic blood pressure (median: 22.79 mmHg) is predicted. These results suggest that AKI-twinX accurately predicts the effects of vasopressors over the next 12 hours.

**Figure 4.**
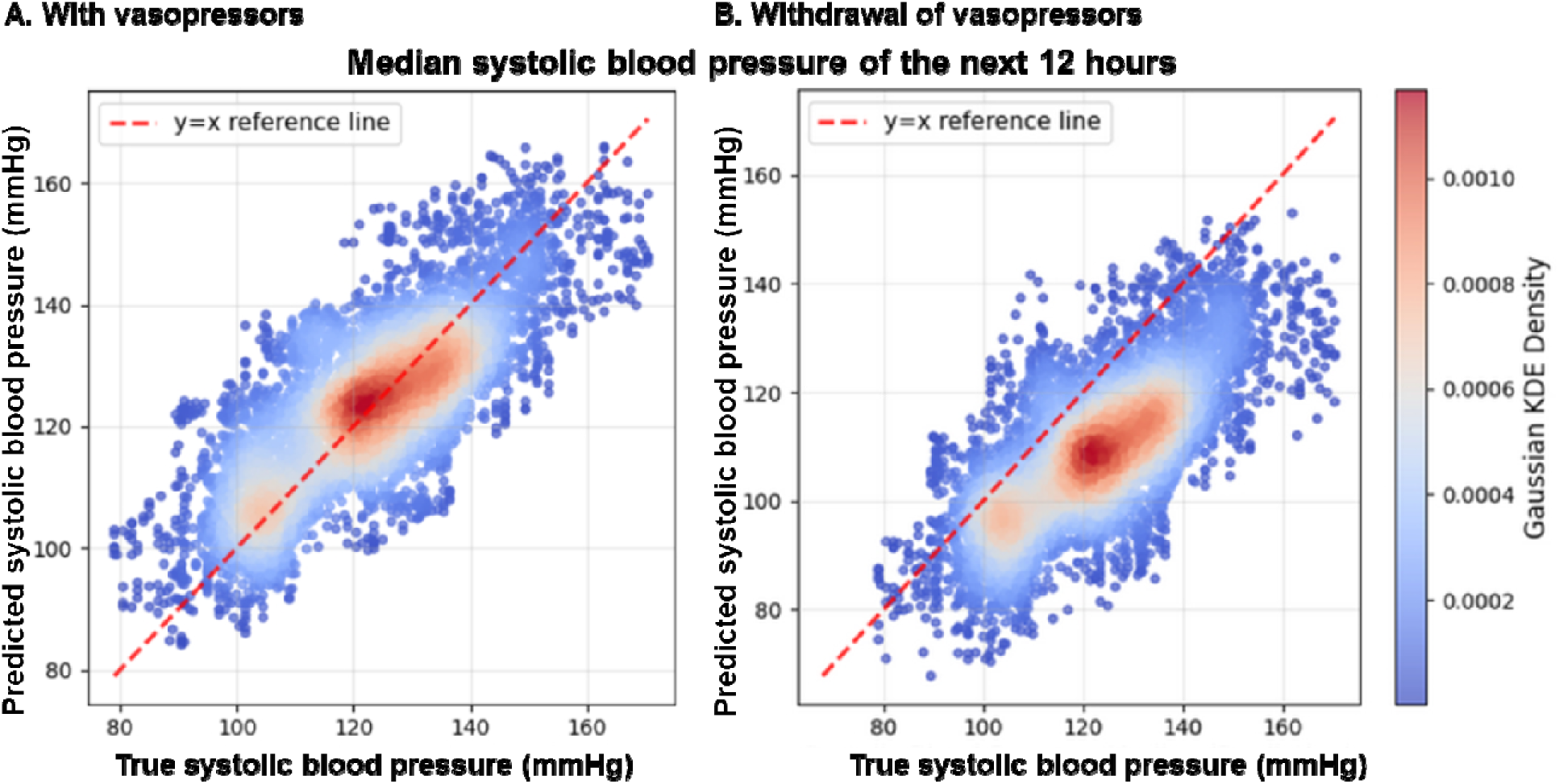
Predicted 12-hour median systolic blood pressure. A. For patients who uses vasopressors. B. For patients who used but assumed the withdraw of vasopressors.

A typical sepsis case in the MIMIC-IV cohort is used to demonstrate a use case for AKI-twinX in an ICU setting (Figure 5). Chart review (Figure 5 A) showed that the patient was an elderly male with sepsis at admission, including urinary tract infection and pneumonia. The patient demonstrated high fever, hypotension, rapid heart rate, acidosis, and elevated white blood cell counts, all suggesting symptoms of septic shock. The patient also had a history of cardiovascular diseases, reduced ejection fraction, hyperlipidemia, and hemiplegia. The patient was under vasopressors (vasopressin and norepinephrine) as well as an antibiotic (vancomycin) at the 47^th^ hour. The design (Figure 5B) is to compare the impact of withdrawing vasopressors. This moment of decision was selected because it marked a pivotal point at which the patient’s clinical condition significantly improved. AKI-twinX suggested that withdrawal of vasopressors significantly increased the risk of death (a 56% increase in the mortality risk from 22.61% to 35.22%, Figure 5 C). The projections of temporal clinical features (Figure 5D) demonstrated that the predicted values (blue lines) are consistent with the true measurements (black lines). The predicted trends after stopping vasopressors would miss the opportunity window of maintaining blood pressure (below 90 mmHg vs above 110 mmHg) and the control of heart rate (above 110 bpm vs below 110 bpm), and stabilizing kidney functions (serum creatinine above 2.25 mg/dL or above 50% increase vs remaining stable below 1.50 mg/dL, and urine output dropped below 0.5 mL/hr vs recovered back to 0.8 mL/hr. The heart status, represented as the predicted cardiovascular SOFA score, remained poor (around 4), but the kidney status would significantly deteriorate (predicted renal SOFA score increased from 1.0 to above 2.0) if vasopressors were stopped, and would improve (predicted renal SOFA score from 1 to below 0.5) if not. This case demonstrates the importance of maintaining vasopressors for this patient at this time and predicts an improvement in clinical condition.

**Figure 5.**
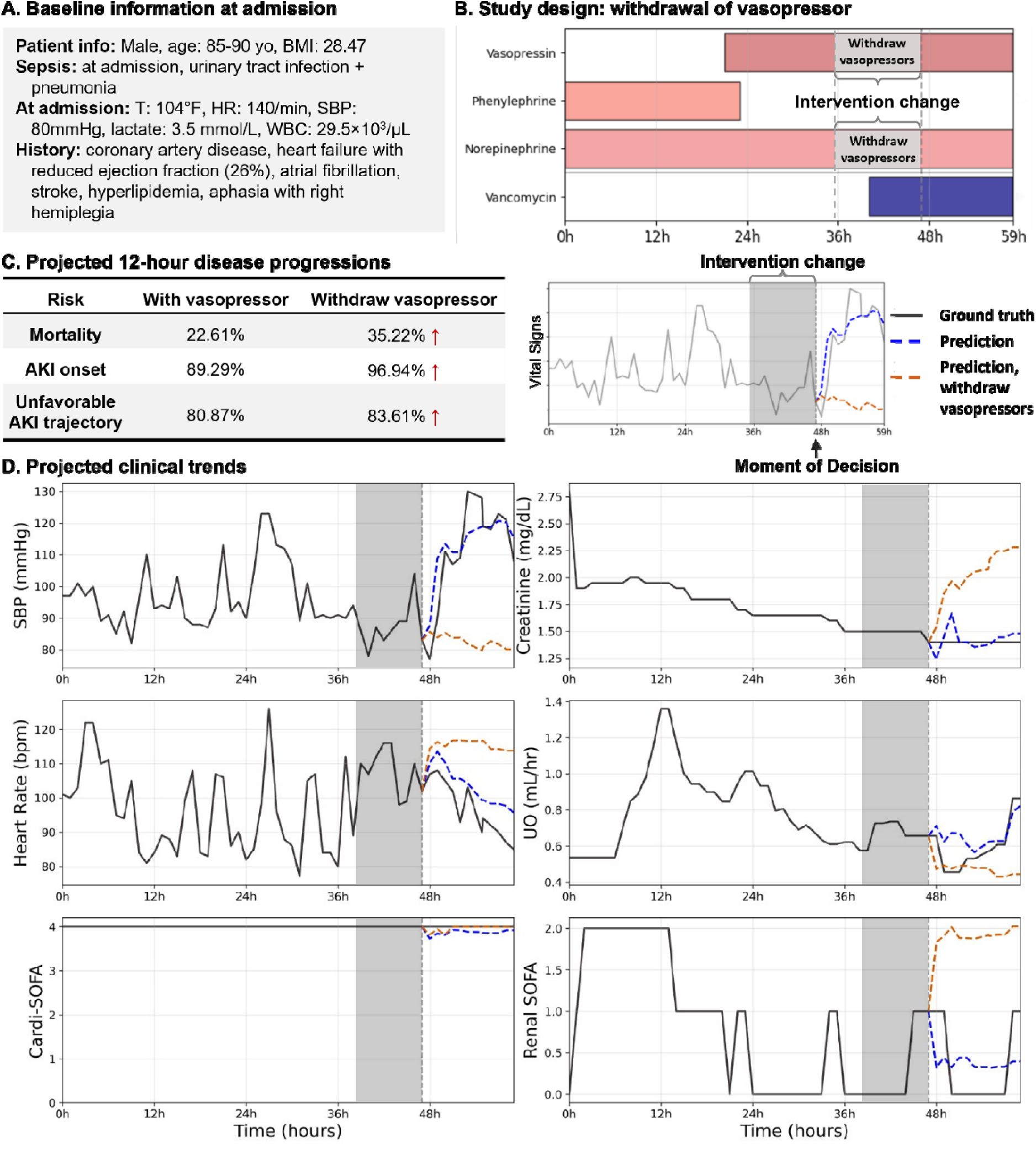
Case study of the selected patient. A. Patient baseline information at admission. B. Illustration of intervention change case study. C. Projected12-hour disease progressions from the model and illustrative guideline. D. Projected clinical trends.

## Discussion

AKI-twinX is an organ-structured intensive care digital twin for sepsis that is designed to support near-term forecasting while keeping the model’s internal reasoning aligned with clinical physiology. The main contribution is the integration of three elements that are rarely combined in a single ICU framework: an explicit organ-level latent structure, multi-task prediction of clinically coupled endpoints, and model-integrated interpretability learned during training rather than applied after the fact. By separating cardiovascular, renal, and other organ representations and then modeling their interactions, AKI-twinX produces internal states that are easier to inspect and debate at the bedside, which is critical in sepsis, where physiologic trajectories evolve quickly, and decisions often hinge on what is likely to happen over the next several hours rather than on a static risk estimate.

A second key observation is that AKI-twinX functions as a short-horizon forward model of physiology while also forecasting outcomes. The model is trained to project near-term trajectories of clinically meaningful variables in parallel with the prediction of survival risk, AKI onset, and AKI trajectory. This matters because ICU clinicians typically do not act solely on the basis of outcome probabilities. They act on whether hemodynamics are likely to remain stable, whether renal function is likely to improve or deteriorate, and whether a proposed change in support will shift the patient into a more fragile state. In your evaluation, the systolic blood pressure projection demonstrates this physiologic fidelity in a measurable way, with predictions closely tracking observed values in vasopressor-treated patients and a coherent separation when vasopressors are counterfactually withdrawn. These properties move the model closer to a clinical “expectation engine” that forecasts what the next window may look like, rather than a classifier that only labels risk.

The framework also addresses an important gap in the AKI prediction literature by shifting the target from AKI occurrence alone to AKI dynamics. Many ICU AKI models focus on detecting or forecasting onset within a fixed horizon, which is useful for early warning but does not fully answer the bedside question once AKI has occurred. After AKI is established, clinicians need to anticipate whether kidney injury is likely to resolve, stabilize, or progress over the next day under ongoing therapies. AKI-twinX explicitly models AKI stage transitions across adjacent windows and then maps those transitions into clinically interpretable trajectory groups, emphasizing the directionality of the renal course rather than a single event. This choice also aligns with sepsis practice, in which renal dysfunction can be reversible and short-term trajectory awareness can inform nephrotoxin exposure management, perfusion targets, and the timing of escalation decisions.

Compared with prior digital twin efforts in healthcare and sepsis, the approach here is distinguished by its balance between physiologic structure and data-driven learning at scale. Mechanistic digital twins and rule-based multi-organ simulators can be interpretable, but they often depend on expert-defined assumptions and have limited external validation. On the other hand, purely data-driven ICU models, including mortality predictors and treatment policy learning systems, can achieve strong predictive performance but often provide limited organ-specific transparency or rely on post hoc explanations that are not stable in temporally correlated ICU data. AKI-twinX takes a different approach by embedding organ-level disentanglement and feature gating directly into the representation-learning process. The learned gating masks yield organ-specific feature-influence patterns that are clinically recognizable, with the renal representation emphasizing systolic blood pressure, short-interval urine output, creatinine-related indices, and renal SOFA, and the cardiovascular representation emphasizing vasoactive and respiratory support, as well as overall severity measures. Importantly, cross-organ interaction modules enable the model to represent cardio-renal coupling without collapsing the physiology into a single, difficult-to-interpret blended embedding.

The clinical utility of this model is best understood as decision support that augments clinician reasoning in three practical settings. First, it enables trajectory-aware monitoring when outcome risks and organ trajectories are discordant, which is common in sepsis. A patient may appear hemodynamically improved while renal vulnerability persists, or renal recovery may lag behind improvements in other systems. Joint forecasting of survival risk, AKI onset, and AKI trajectory can help teams identify discordant patterns early and prioritize clinically standard but often delayed actions, including targeted review of nephrotoxic exposures, reassessment of perfusion adequacy, and earlier nephrology engagement when the predicted renal course remains unfavorable. Second, the model supports structured what-if reasoning for vasoactive management, which is a frequent and consequential decision point in septic shock. The case illustration is clinically representative: an elderly patient with severe cardiac dysfunction shows apparent improvement, yet remains physiologically dependent on vasopressors. In that context, the digital twin’s projection under continued support versus simulated withdrawal provides an explicit forecast of hemodynamic instability, renal deterioration, and increased downstream risk, offering a safety-oriented lens for tapering decisions rather than an automated recommendation. Third, model-integrated interpretability supports operational trust. When predictions disagree with bedside impressions, clinicians can examine whether the forecast is being driven by plausible organ-specific signals or by artifacts such as documentation gaps, stale laboratory carry-forward, or exposure misclassification, which is particularly important for real-world ICU deployment.

Transportability is a major barrier to ICU AI, and external validation across independent health systems strengthens the readiness argument for this framework. The model was developed in a large public ICU dataset and then validated in a separate multi-hospital cohort with different data provenance and documentation patterns, yet maintained consistent discrimination across the major prediction tasks. This cross-system consistency supports the idea that organ-structured representation learning can reduce overfitting to institution-specific workflows. At the same time, external validation should be treated as the start of implementation, not the end. The next step is a prospective, silent deployment that tracks calibration, stability, and input drift over time, followed by a clinician-in-the-loop pilot that evaluates whether the tool improves decision-making and workflow rather than merely predictive performance.

Several limitations warrant emphasis. Most importantly, the counterfactual intervention analyses are not causal estimates. Vasopressor continuation and withdrawal are confounded by clinician intent, evolving severity, and unmeasured physiology; therefore, the simulated withdrawal results should be interpreted as model-based forecasts under altered inputs rather than as treatment effects. In addition, intervention representation is simplified. Vasopressor exposures are encoded without fully capturing dose titration dynamics, individualized targets, and clinician response loops, and other major levers in sepsis care, including fluid strategy, ventilation settings, renal replacement therapy initiation, and antimicrobial adjustments, are not modeled with comparable granularity. Data harmonization constraints also matter, particularly the incomplete stop-time documentation in the IUH cohort and the resulting exposure-persistence assumptions, which can introduce misclassification and affect both feature-importance patterns and counterfactual outputs. Outcome definitions impose additional constraints: grouping AKI stage transitions into favorable versus unfavorable trajectories improves interpretability but can obscure clinically meaningful heterogeneity within each group, and SOFA-based organ labels are widely used yet remain imperfect proxies that reflect both physiology and care processes.

Finally, as with any retrospective EHR model, AKI-twinX is vulnerable to temporal shift. Documentation practices, order sets, vasopressor selection patterns, and laboratory cadence can change, potentially degrading calibration even when discrimination remains stable. This risk is particularly relevant for models intended for real-time use. Subgroup robustness also requires explicit evaluation because sepsis severity, baseline comorbidity, and treatment intensity may differ across age, sex, race, and ethnicity, and ICU types. These limitations point toward concrete next steps, including drift monitoring with predefined recalibration triggers, systematic subgroup audits, and a prospective evaluation focused on clinical impact, interpretability, and human factors rather than performance metrics alone.

In summary, AKI-twinX supports the feasibility of an organ-structured intensive care digital twin that jointly forecasts survival risk, AKI onset, and AKI trajectory while providing organ-aligned interpretability and physiologically coherent short-horizon projections. The distinctive value is not a single performance number, but a model design that aligns representation learning with organ reasoning, supports clinically meaningful simulation around vasoactive management, and remains consistent across independent cohorts. Prospective silent deployment with calibration and drift monitoring, richer intervention modeling that incorporates dose and timing, and workflow-focused human factors studies are the most appropriate next steps toward safe and practical ICU implementation.

## Methods

### Datasets

We used two large critical care databases: the Medical Information Mart for Intensive Care IV (MIMIC-IV, v2.2) and the Indiana University Health (IUH) ICU dataset (v1.0). MIMIC-IV includes 73,181 ICU stays from Beth Israel Deaconess Medical Center (Boston, Massachusetts, USA) recorded between 2008 and 2019. The IUH dataset includes 6,918 ICU admissions across 11 hospitals in Indiana, USA, from 2019 to 2023. Dataset composition and preprocessing are summarized in Supplementary Fig. 1A and 1B.

We included ICU admissions from adults (age ≥18 years) with heart rate and blood pressure measurements and documented sex, race, and survival status. To ensure adequate temporal coverage for trajectory modeling, we limited the analysis to ICU stays of at least 48 hours. We further restricted the cohort to admissions with a confirmed diagnosis of sepsis to focus on cardio-renal interactions during critical infection. Each ICU admission was treated as a separate analytic unit, even when multiple admissions occurred in the same patient. This reflects the episodic nature of critical illness and potential changes in management across stays. Cohort characteristics are reported in Supplementary Table 1.

### Input features

Predictors included baseline characteristics measured at ICU admission and time-varying clinical features derived from the 12-hour window preceding each prediction (Supplementary Table 2). Baseline features comprised demographics (age, sex, height, weight, and body mass index) and comorbidity burden, represented by the Charlson Comorbidity Index, including the composite score and individual components (e.g., myocardial infarction, congestive heart failure, chronic pulmonary disease, diabetes, renal disease, and malignancy). The baseline feature set also included the most recent renal and cardiovascular SOFA subscores available within the 12-hour window.

Time-varying features were extracted from the 12-hour prediction window and represented as a sequence of 12 time steps, using hourly bins after preprocessing. These features included vital signs (temperature, available blood pressure measurements, and respiratory rate), SOFA subscores for coagulation, liver, cardiovascular, central nervous system, and renal function, as well as the total SOFA score. We also included renal and metabolic markers, including blood urea nitrogen, serum creatinine, the blood urea nitrogen–to–creatinine ratio, platelet count, bicarbonate, and free calcium. Urine output rates were computed over 6-, 12-, and 24-hour intervals. Indicators of respiratory support included invasive and non-invasive ventilation and oxygen therapy, and vasoactive exposure included norepinephrine, dopamine, dobutamine, epinephrine, milrinone, phenylephrine, and vasopressin.

Medication exposures during the observation window were incorporated as additional predictors. These included antibiotic categories and selected agents, cardiovascular medications, and other clinically relevant exposures related to renal vulnerability, including proton pump inhibitors, contrast agents, and nonsteroidal anti-inflammatory drugs. Drug categories and included medications are listed in Supplementary Table 3.

### Preprocessing

We applied prespecified exclusion criteria during cohort construction to ensure adequate demographic annotation and physiologic coverage. ICU stays missing age, sex, race, survival status, or heart rate and blood pressure measurements were excluded (Supplementary Fig. 1A and 1B). Time-varying variables were standardized to an hourly resolution, and all features were aligned to a common timeline.

Missing data were handled using feature-specific rules. Baseline variables with partial missingness, including height, weight, and body mass index, were imputed using the cohort median. SOFA subscores were forward-filled from the most recent observation. Respiratory support indicators were coded as absent when documentation was unavailable. We did not impute entire missing feature groups. ICU stays with a complete absence of key groups, such as SOFA data or respiratory support documentation, were excluded rather than imputed.

Medication exposures were aligned to the hourly timeline by propagating initiation times through documented discontinuation. In the IUH dataset, where stop times were inconsistently recorded, we applied a 12-hour decay window after each administration. Laboratory measurements were aligned to each prediction window by carrying forward the closest available value. When two measurements flanked a window, we interpolated using the median of the two values. ICU stays with a complete absence of heart rate or systolic blood pressure within a prediction window were excluded. For partially missing heart rate and systolic blood pressure within a window, we did not impute missing points and summarized these signals at the window level using median values. We applied the same approach to other time-varying clinical features.

### Phase 1: Generative latent-state model for short-horizon forecasting

The first stage of AKI-twinX is a generative latent-state model that learns organ-structured representations of patient trajectories and supports short-horizon forecasting (Supplementary Fig. 2A). At each prediction step, the model takes two inputs. The first is a 12-hour sequence of time-varying clinical features, represented as a 12-hour-by-49-variable matrix. The second is a baseline feature vector with 42 variables that captures demographics and comorbidities and includes the most recent renal and cardiovascular SOFA subscores available within the same 12-hour window. This design combines short-term physiologic context with baseline risk information while maintaining a consistent time window across model stages.

To encode organ-specific dynamics, we used three parallel variational encoder branches aligned to cardiovascular, renal, and other organ function. Each branch begins with a gated recurrent unit that summarizes temporal dependencies in the 12-hour sequence. The summarized representation is then mapped to a Gaussian latent distribution parameterized by a mean vector and a log-variance vector. We sample latent vectors using the reparameterization trick, with an isotropic standard normal prior. The cardiovascular and renal encoders are front-gated by learnable feature masks with L1 regularization. These masks encourage sparse feature use and yield organ-specific feature rankings that can be inspected. The “other organ” encoder does not use feature selection and is intended to capture remaining shared and non-cardio-renal signals. The three encoders output organ-aligned latent vectors for cardiovascular, renal, and other organ channels. We also apply an orthogonality constraint across these latent subspaces to reduce redundancy and promote disentanglement, ensuring that each subspace preferentially captures information specific to its target organ system.

Baseline information is integrated through three organ-aligned feed-forward baseline processors. Feature gating is applied to the cardiovascular and renal baseline processors to align with the temporal encoders and preserve the interpretability of baseline inputs. Each baseline embedding is concatenated with its corresponding temporal latent vector, producing an enriched organ-specific representation that combines baseline context with recent dynamics.

The model uses three decoders to generate short-horizon forecasts from the latent space. The cardiovascular decoder predicts the cardiovascular SOFA signal for the next 12-hour window, and the renal decoder predicts the renal SOFA signal for the next 12-hour window. The third decoder forecasts the time-varying clinical feature sequence over the subsequent 12 hours. This is a forward prediction objective rather than a same-window reconstruction objective. It encourages the latent space to encode dynamics that are informative for both physiologic trajectory forecasting and organ status evolution.

Training optimizes a composite objective that balances forecasting accuracy, organ supervision, interpretability, and disentanglement. We use mean squared error losses for next-window feature sequence forecasting and for cardiovascular and renal SOFA regressions. We apply L1 regularization to the gating masks to promote sparsity. We include a Kullback–Leibler divergence term for variational training and an orthogonality loss to encourage separation of the organ-specific latent subspaces. To improve stability, we use weighted loss scheduling across epochs. Early training emphasizes learning core temporal dynamics, whereas later training places greater emphasis on organ separation and feature sparsity objectives.

### Phase 2: Outcome and trajectory prediction model

The second stage of AKI-twinX is a prediction model that uses the organ-specific latent representations learned in Phase 1 to forecast near-term outcomes and AKI dynamics (Supplementary Fig. 2B). At each moment of prediction, we reused the Phase 1 temporal encoders and baseline processors to generate latent vectors for the cardiovascular, renal, and other-organ channels. To preserve the organ-aligned structure learned during generative training, all Phase 1 components were kept frozen during Phase 2. For each organ channel, we concatenated the temporal latent vector with its baseline embedding to form an enriched, organ-specific representation that integrates recent physiological information with baseline context.

Because sepsis is characterized by strong cross-organ coupling, we modeled interactions across the three organ representations using a cross-attention module. This mechanism allows each organ channel to condition on the others, capturing cardio-renal and broader multi-organ dependencies without collapsing the representation into a single undifferentiated embedding. The attention-augmented organ vectors were then fused and passed to three task-specific prediction heads.

The mortality head predicted survival status over the next 12-hour window. The AKI onset head predicted incident AKI in the next 12-hour window, defined as new AKI stage 1 or higher among admissions without AKI in the immediately preceding 12-hour window. The AKI trajectory head model was trained to predict changes in AKI stage across adjacent 12-hour windows and produced three clinically interpretable categories that summarize the short-horizon renal course. To reflect the ordinal structure of AKI severity, the trajectory head used a threshold-based ordinal formulation. It learned a continuous latent severity score and mapped it to favorable, stable, or unfavorable trajectory classes using two learned thresholds, avoiding an unstructured categorical treatment of stage progression.

Phase 2 training jointly optimized the three prediction heads. The mortality and AKI onset heads used binary cross-entropy loss, and the trajectory head used the ordinal loss described above. Task-specific loss weights were selected based on validation performance to balance multi-task learning, while the frozen Phase 1 encoders preserved the organ-specific latent spaces learned during Phase 1.

### Model training, cross-validation, and evaluation

Model development and internal evaluation employed five-fold cross-validation on the MIMIC-IV cohort, followed by external validation on the IUH sepsis cohort. Training proceeded sequentially, with Phase 1 (the generative latent-state model) trained first and Phase 2 (the prediction model) trained on frozen Phase 1 encoders. Data splits were performed at the ICU-stay level, and all prediction windows from a given ICU stay were assigned to the same fold to prevent leakage across folds.

For Phase 1, we used the Adam optimizer with an initial learning rate of 1X10^−5^. After 75 epochs, the learning rate was increased to 1X10^−3^ to accelerate learning of temporal patterns. Training ran for up to 200 epochs with a batch size of 256. We applied early stopping when the validation mean squared error failed to improve by at least 0.001 over 10 consecutive epochs. To stabilize optimization across the multiple objectives, we used a weighted loss schedule with stepwise reweighting over epochs. At initialization, the kidney SOFA loss was emphasized (kidney SOFA weight 1.0), with next-window feature forecasting and heart SOFA losses set to zero, KL weight set to 0.1, L1 weight set to 0.01, and the orthogonality loss set to zero. At epoch 40, we activated the heart SOFA and orthogonality losses (heart SOFA weight 1.0, kidney SOFA weight 1.0, orthogonality weight 0.1) while keeping next-window feature forecasting inactive. At epoch 75, we activated the next-window feature forecasting and increased the emphasis on SOFA prediction (forecasting weight 1.0, heart SOFA weight 10.0, kidney SOFA weight 10.0) while maintaining the KL, L1, and orthogonality weights.

For Phase 2, we trained prediction heads using Adam with a learning rate of 1X10^−3^ and weight decay of 1X10^−4^. We applied a ReduceLROnPlateau scheduler that halved the learning rate when the validation AUC did not improve for 10 epochs. Training ran for up to 100 epochs with early stopping when the validation AUC failed to improve by at least 0.001 over 10 consecutive epochs, using the mean AUC across the mortality and AKI onset tasks as the stopping criterion. Loss functions were binary cross-entropy for mortality and AKI onset, and the ordinal loss described above for AKI trajectory prediction.

We monitored Phase 1 using mean squared error for next-window feature forecasting and SOFA score prediction. Phase 2 performance was evaluated primarily using AUC, including binary AUC for mortality and AKI onset. For the AKI trajectory, we report macro-averaged AUC and the concordance index.

### Case-based intervention illustration

To illustrate the potential clinical utility of AKI-twinX, we selected a representative admission from the MIMIC-IV sepsis cohort. The patient was a male aged 85-89 years with substantial cardiovascular comorbidity, including coronary artery disease, heart failure with reduced ejection fraction (26%), atrial fibrillation, and prior ischemic stroke. He was admitted from a nursing home with septic shock attributed to urinary tract infection and pneumonia, complicated by hypoxemia, hypotension, tachyarrhythmia, and acute kidney injury. Management included intubation, broad-spectrum antibiotics, and vasopressor support with norepinephrine and phenylephrine.

For this admission, the model generated 12-hour forecasts of heart rate, systolic blood pressure, creatinine, urine output (6-hour rate), and cardiovascular and renal SOFA subscores, along with predictions of AKI onset risk, AKI trajectory, and short-horizon mortality risk. We then performed a counterfactual simulation in which norepinephrine and phenylephrine were set to absent throughout the preceding 12-hour window and recomputed the same forecasts for comparison. This counterfactual analysis is intended as a model-based sensitivity probe and does not estimate a treatment effect.

Under observed therapy, the model forecasts tracked the measured trajectories across the physiologic variables and SOFA subscores. Under simulated vasopressor withdrawal, the recomputed forecasts shifted in clinically coherent directions, with higher heart rate, lower systolic blood pressure, worsening renal markers, lower urine output, and higher cardiovascular and renal SOFA signals, accompanied by higher predicted risks of AKI onset and mortality. This case reflects a common bedside scenario in septic shock, in which an older patient with limited cardiac reserve may appear improved yet remain dependent on vasoactive support. It illustrates how a digital twin can be used to compare projected short-horizon trajectories under alternative support patterns.

### Short-horizon systolic blood pressure forecasting and counterfactual vasopressor withdrawal

To assess physiologic fidelity, we evaluated the model’s ability to forecast systolic blood pressure over the next 12 hours. Using a representative model trained on the MIMIC-IV cohort, we analyzed 6,673 test samples. Predicted median systolic blood pressure values closely tracked observed medians across the measurement range, with points aligning near the identity line (Fig. 4A). We then examined sensitivity to vasoactive support using a counterfactual withdrawal analysis by setting norepinephrine, phenylephrine, vasopressin, and epinephrine absent throughout the 12 hours preceding prediction and recomputing systolic blood pressure forecasts. Relative to predictions under observed treatment, the counterfactual distributions shifted away from the identity line and formed a clear angular separation (Fig. 4B), reflecting lower predicted systolic blood pressure under simulated withdrawal. This counterfactual comparison is intended as a model-based sensitivity probe and does not estimate a treatment effect.

### Organ-specific feature importance from learned gating masks

In Phase 1, we applied learnable gating masks to baseline and time-varying inputs to encourage sparse feature use within the cardiovascular and renal encoders. We then summarized feature importance using the learned mask weights to identify inputs that most strongly contributed to each organ branch (Fig. 3). The cardiovascular branch was dominated by signals reflecting respiratory support and vasoactive therapy, whereas the renal branch emphasized systolic blood pressure, creatinine, urine output, and medication exposures relevant to renal vulnerability. This organ-level decomposition links latent states to recognizable clinical inputs and provides transparency into how the model separates cardio-renal dynamics.

## Declarations

Indiana University Human Subject & Institutional Review Boards ethical review granted an IRB waiver for the project.

## Data availability

The IUH dataset is available via email to Lingzhong Meng and is pending institutional approval. The MIMIC-IV datasets are available from their respective sources.

## Code availability

The code used in the current study to develop the algorithm will be made available on GitHub.

## Acknowledgements

We thank Rick V. Tuason from Clinical Research Systems, Enterprise Analytics, Indiana University Health, located in Indianapolis, Indiana, USA, for his help in preparing the Indiana University Health dataset. J.S. and A.E.G. were financially supported by the National Library of Medicine of the National Institutes of Health under award number R01LM013771. J.S. and J.C. were financially supported by the Indiana University Precision Health Initiative. We also thank institutional and/or departmental sources for their support.

## Author contributions

J.S., L.M. and B.Y. conceived and supervised the project. J.S., J.C., and B.Y. designed the model and computational framework. J.S., L.M., M.T.E., and A.E.G. designed the study. J.L., D.P., and A.E.G. collected and prepared data. J.C. performed the statistical analysis. All authors provided critical comments and reviewed the paper. They discussed the results and approved the final version before submission.

## Additional information

Supplementary materials

Correspondence and material requests should be addressed to Jing Su, Lingzhong Meng, or Baijian Yang.

