## Supplement for "AKI-twinX: explainable organ structured digital twin for sepsis AKI trajectory forecasting"

The authors provide the supplementary materials for the following manuscript:

**“AKI-twinX: an explainable intensive care digital twin for managing AKI progression”**

**Content**:

- **Supplementary Figure 1a:** Cohort consort diagram for MIMIC IV data
- **Supplementary Figure 1b**: Cohort consort diagram for IUH data
- **Supplementary Table 1**: Characteristics of the two datasets used in this study
- **Supplementary Table 2**: Features included in the model
- **Supplementary Table 3**: Categories of drugs included in feature model and the medications included in these categories
- **Supplementary Figure 2**: Diagram explaining model architecture

**Supplementary Figure 1a:** Cohort consort diagram for MIMIC IV data

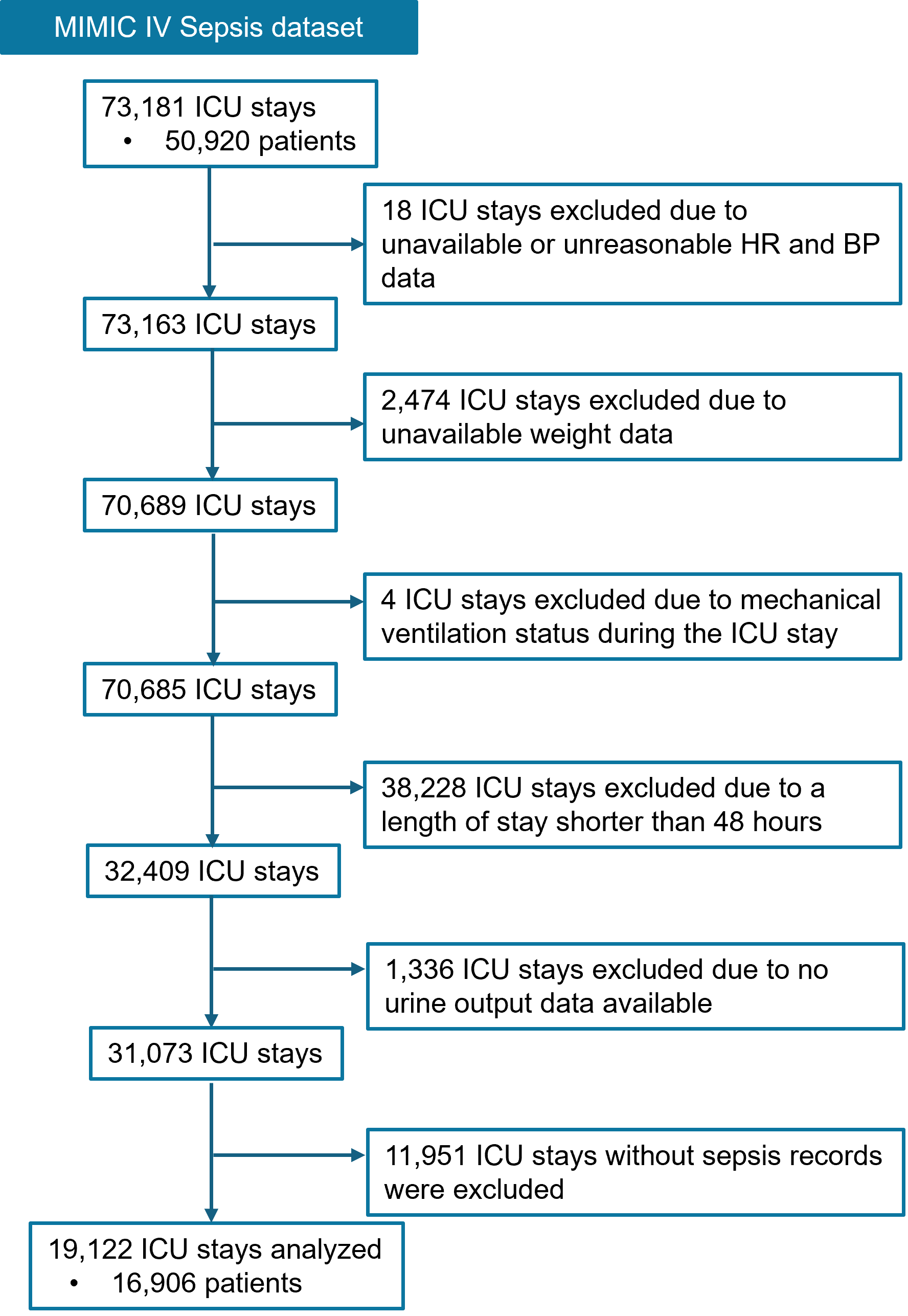

ICU, intensive care unit

**Supplementary Figure 1b**: Cohort consort diagram for IUH data

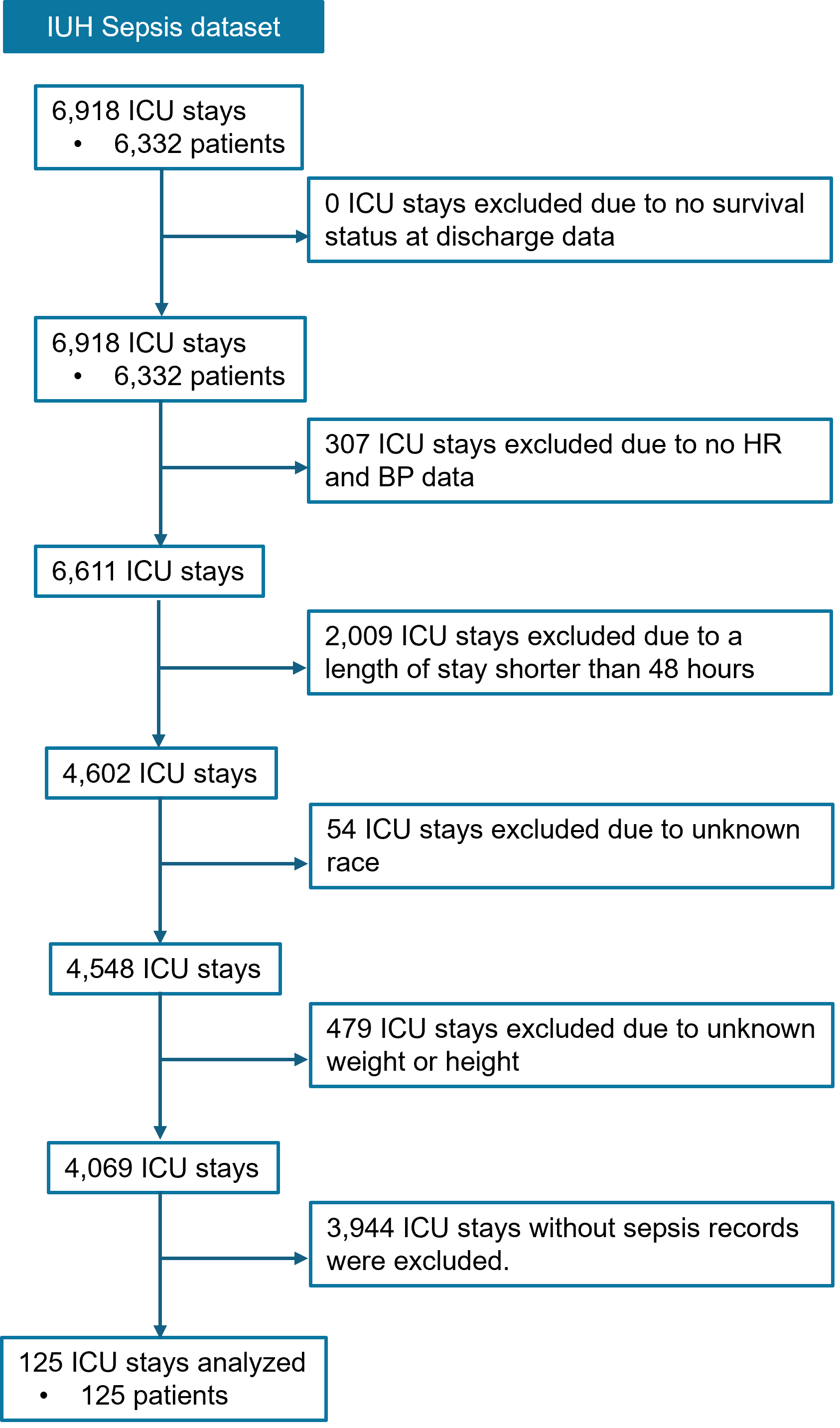

IUH, Indiana University Health; ICU, intensive care unit

**Supplementary Table 1**: Characteristics of the two datasets used in this study

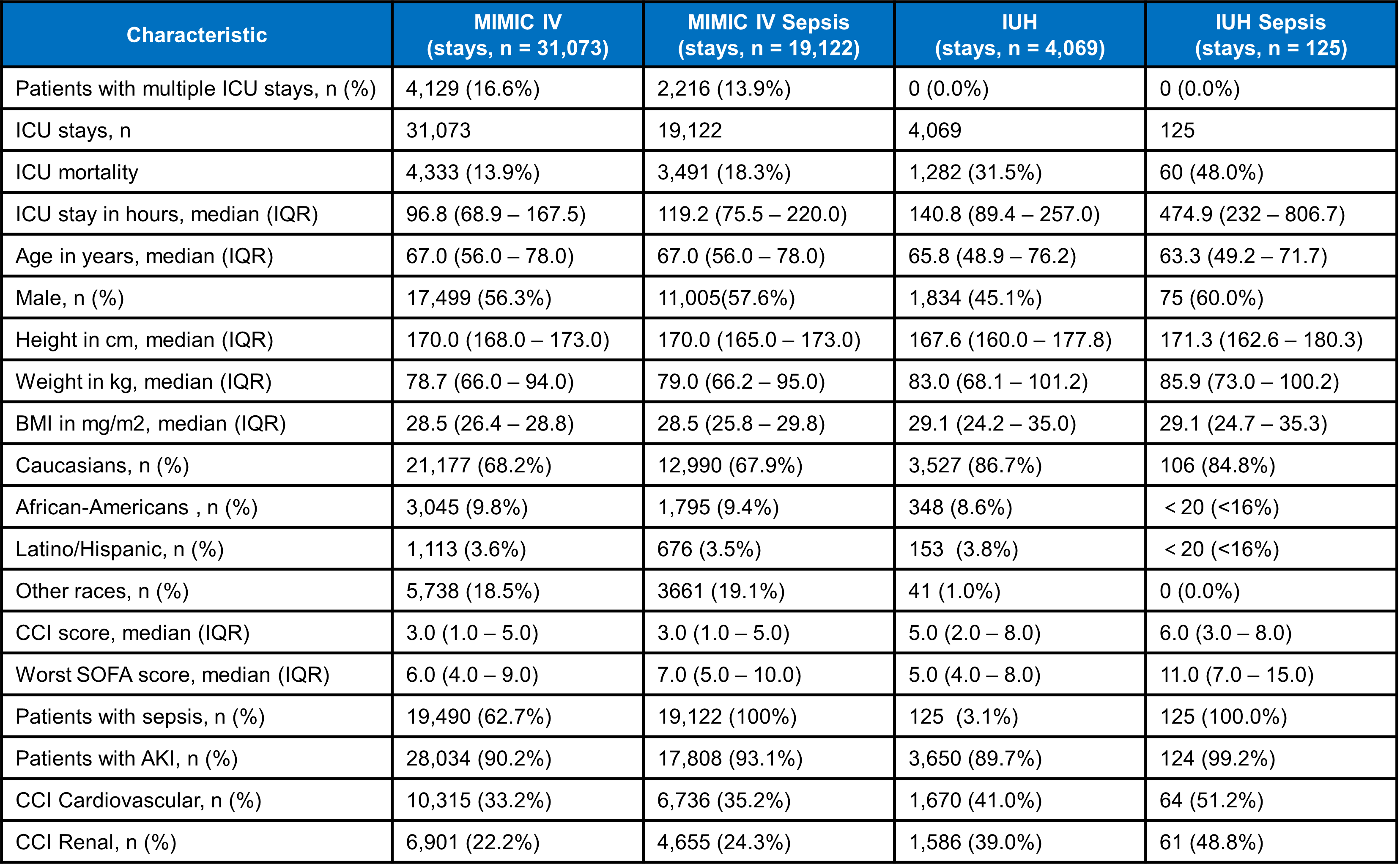

IUH, Indiana University Health; ICU, intensive care unit; IQR, interquartile range; BMI, body mass index; CCI, Charles Comorbidity Index; SOFA, Sequential Organ Failure Assessment

Note: ICU stays refer to all ICU admissions recorded in the dataset, each with a unique admission and discharge/mortality date. A single patient may have multiple ICU stays. ICU stays were excluded from the datasets if they had unknown survival status, missing HR or BP data, age below 18 years, or unspecified sex (see Supplementary Figure 1). The IUH dataset includes ICU stays with admission dates from 1 January 2022 onward. ICU mortality refers to death occurring during an ICU stay. For patients with multiple hospitalizations, only the most recent hospitalization was counted. If a patient had multiple ICU stays, calculations were based on data from the last stay. The worst SOFA score is the highest score within the first 24 hours after ICU admission.

**Supplementary Table 2**: Features included in the model

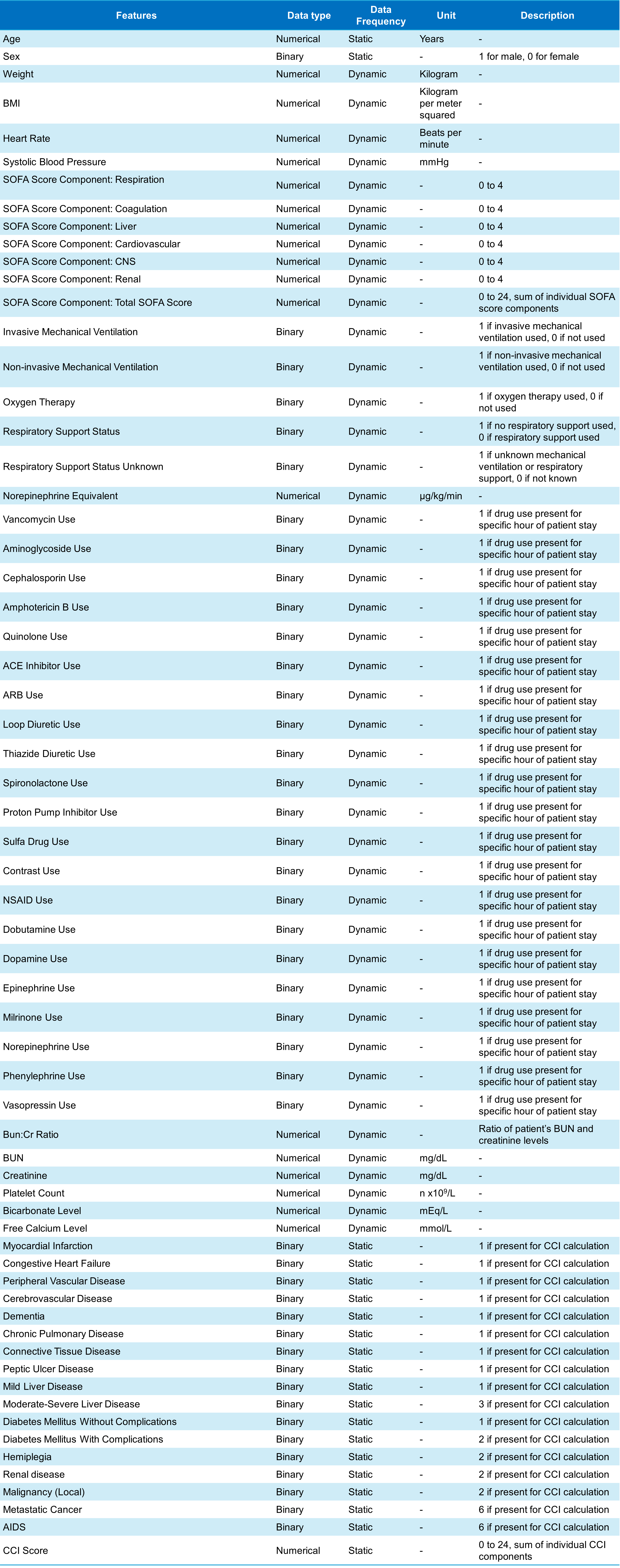

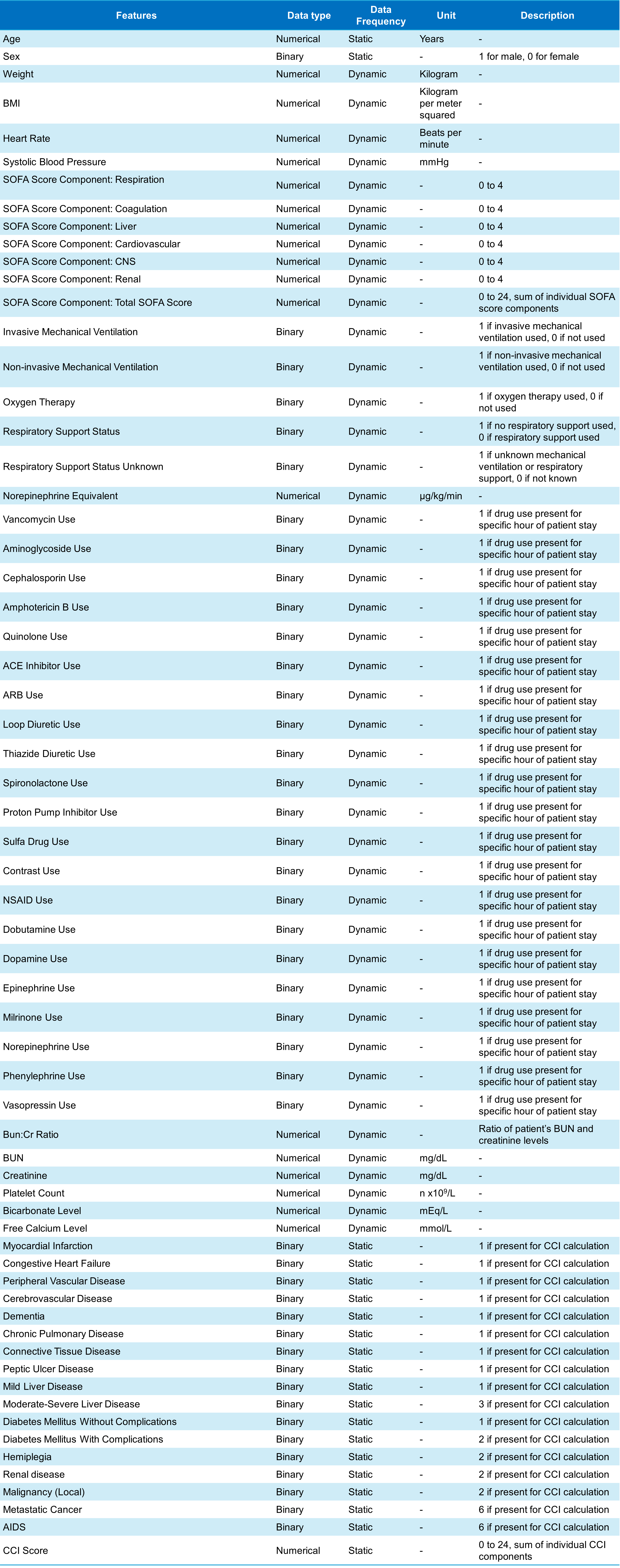

BMI, body mass index; SOFA, Sequential Organ Failure Assessment; Cr, serum creatinine; BUN, blood urea nitrogen; CCI, Charles Comorbidity Index.

The Charlson Comorbidity Index was calculated based on the following conditions: myocardial infarction (1 point), congestive heart failure (1 point), peripheral vascular disease (1 point), cerebrovascular disease (1 point), dementia (1 point), chronic pulmonary disease (1 point), rheumatic disease (1 point), peptic ulcer disease (1 point), mild liver disease (1 point), diabetes without chronic complications (1 point), diabetes with chronic complications (2 points), hemiplegia or paraplegia (2 points), renal disease (2 points), any malignancy (including leukemia and lymphoma, 2 points), moderate or severe liver disease (3 points), metastatic solid tumor (6 points), and HIV/AIDS (6 points). The model incorporated both the presence of each comorbidity (binary) and the total score (numerical).

**Supplementary Table 3**: Categories of drugs included in the feature model and the medications included in these categories

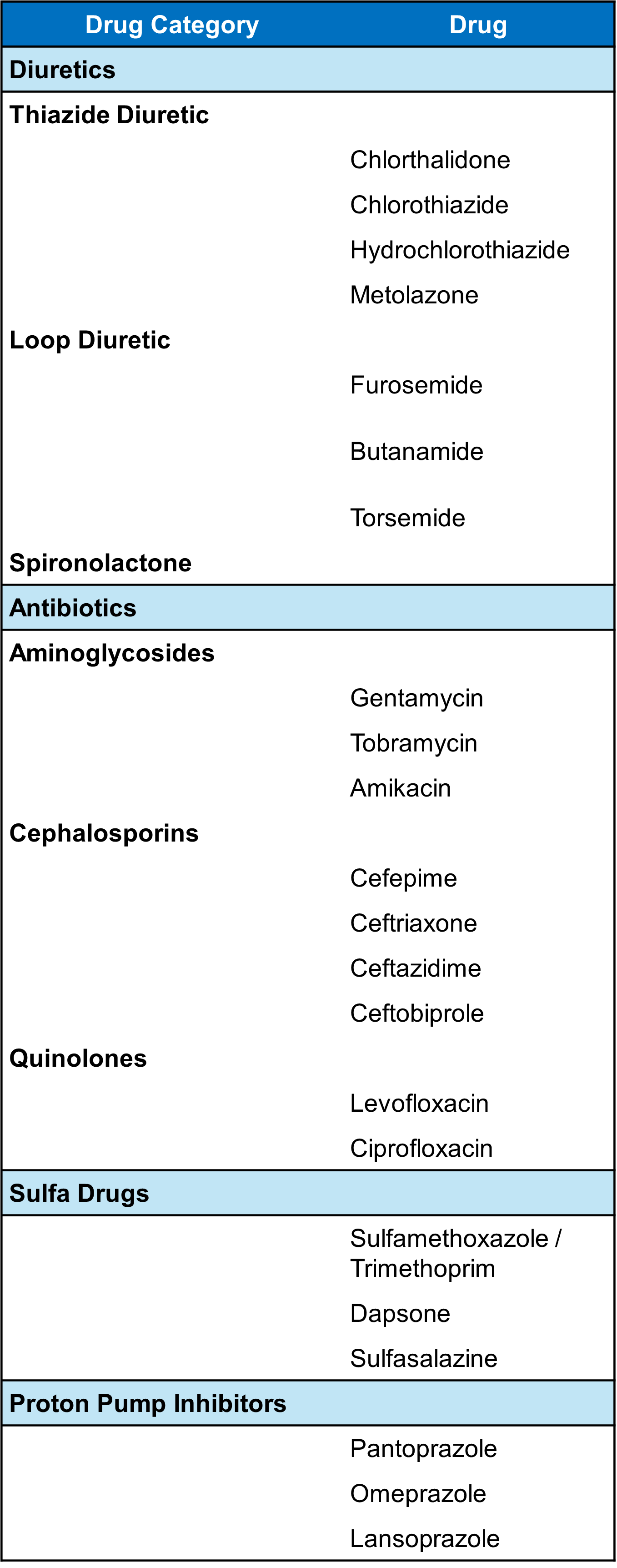

**Supplementary Figure 2**: Diagram explaining model architecture.

(A) Phase I: generative model

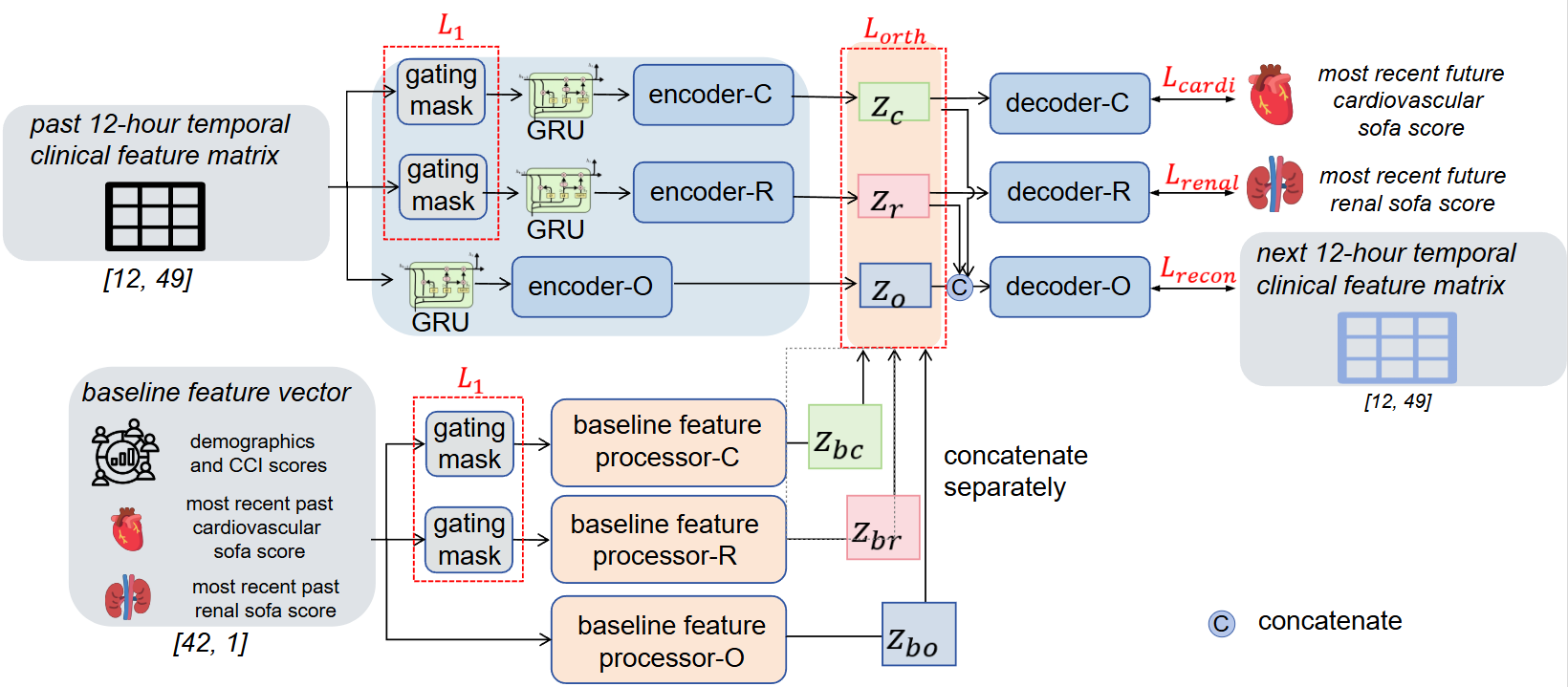

(B) Phase II: prediction model

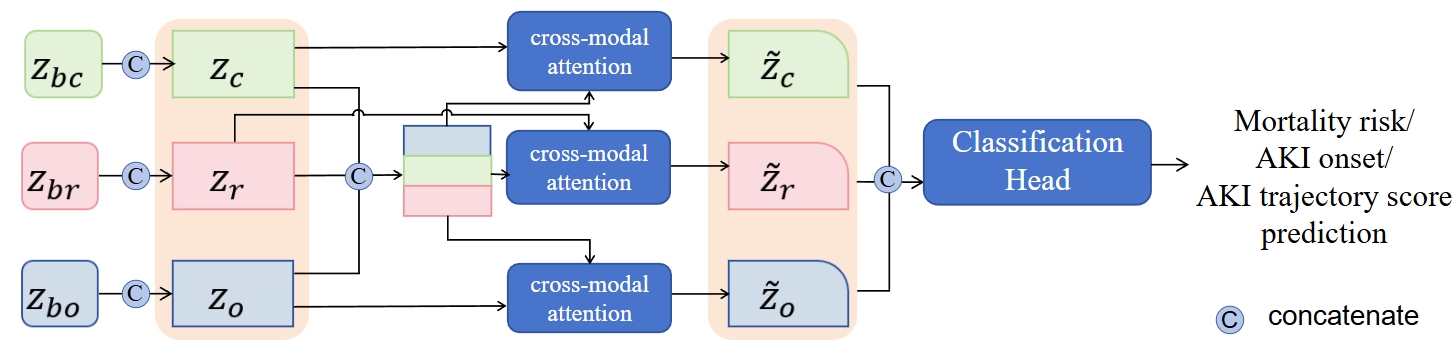

GRU, Gated Recruitment Unit; AKI, acute kidney injury.
